# Operative Hysteroscopy in the Management of Endometrial Polyps: Clinical Indications and Surgical Outcomes

**DOI:** 10.64898/2026.03.31.26349904

**Authors:** Teymur Bornaun

## Abstract

**Background/Objectives:** This study aimed to evaluate the efficacy and outcomes of operative hysteroscopy for the removal of endometrial polyps and assess the procedure’s impact on pain experienced by patients. The research was conducted to determine whether the minimally invasive nature of operative hysteroscopy compromises patient comfort when compared with diagnostic hysteroscopy.

**Methods:** The study was conducted at the Gynecology and Obstetrics Clinic of Bağcilar Training and Research Hospital over a period of four months. It included 200 women over 18 years of age who were indicated for hysteroscopy. Operative hysteroscopy procedures were performed without the use of a speculum, cervical dilation, anesthesia, or analgesic agents, emphasizing the procedure’s minimally invasive approach. Pain assessment utilized the Visual Analog Scale (VAS). Patients were stratified into two groups—those undergoing operative and those undergoing diagnostic hysteroscopy—to compare outcomes and pain scores.

**Results:** The study found that operative hysteroscopy successfully removed 85.1% of the lesions, primarily polyps. There was no significant difference in pain scores between the operative and diagnostic hysteroscopy groups, indicating that the minimally invasive procedure does not increase patient discomfort.

**Conclusions:** Operative hysteroscopy is an effective and tolerable procedure for the removal of endometrial polyps, with high success in complete lesion removal and without significantly impacting the pain experienced by patients. The findings support the use of operative hysteroscopy as a first-line treatment option for endometrial polyps, underscoring the importance of patient selection and the need for further studies on long-term outcomes related to fertility and recurrence.

## 1. Introduction

Endometrial polyps are pathologically benign entities that represent localized hyperplastic proliferations of glandular and stromal components of the endometrium. Despite their benign nature, they are significantly impactful on women’s reproductive health, manifesting through symptoms such as abnormal uterine bleeding, pain, and playing a notable role in infertility [1]. These polyps are among the most common gynecological abnormalities encountered in clinical practice, affecting a wide demographic range, from premenopausal to postmenopausal women.

The clinical management of endometrial polyps has evolved significantly, with a shift towards more conservative and less invasive treatment modalities. Operative hysteroscopy has emerged as a critical innovation in this field, providing a direct visual assessment of the uterine cavity that allows for precise surgical interventions. This technique not only facilitates the removal of polyps but also minimizes patient discomfort and recovery time when compared to traditional surgical approaches such as dilation and curettage [2].

Advancements in hysteroscopic technology, including the development of smaller, more flexible instruments and enhanced imaging capabilities, have significantly broadened the applicability of this procedure. These technological improvements have enhanced the safety profile of operative hysteroscopy, making it a preferred method in the therapeutic arsenal against endometrial polyps [3]. As the equipment and techniques have evolved, so too has the understanding of the procedure’s indications, which now encompass a range of clinical scenarios from symptomatic cases presenting with severe symptoms to asymptomatic cases identified during routine examinations [4].

This study explores the multifaceted role of operative hysteroscopy in the management of endometrial polyps, examining its efficacy and outcomes in both eliminating the polyps and addressing the associated symptoms. It aims to provide a comprehensive overview of current practices and recent advances in the field, highlighting the procedure’s impact on improving patient outcomes and its integration into standard gynecological care.

Operative hysteroscopy offers a dual advantage of diagnosis and treatment, providing a minimally invasive option that can be performed under direct visualization. This not only ensures the thorough removal of polyps but also allows for immediate histopathological evaluation. Such capabilities are crucial in distinguishing benign from potentially malignant polyps, especially in cases where there are atypical presentations or higher risks such as in postmenopausal women or those with a history of hormonal therapy [5].

The procedure’s versatility extends to its diagnostic capabilities, where it serves as a gold standard for the direct assessment of the uterine cavity, allowing clinicians to make informed decisions based on real-time visual evidence. This level of precision is essential for tailoring individual treatment plans and for increasing the overall effectiveness of interventions aimed at reducing symptoms and improving reproductive outcomes.

Furthermore, this review will delve into the socio-economic implications of adopting operative hysteroscopy on a wider scale, considering aspects such as cost-effectiveness, patient satisfaction, and its potential to reduce the overall burden on healthcare systems. By examining a variety of case studies and pooling data from numerous clinical trials, this document will synthesize key findings to offer a nuanced perspective on the role of operative hysteroscopy in modern gynecology.

## 2. Materials and Methods

### 2.1. Study Design

This prospective cohort study was conducted to assess the outcomes and pain associated with operative and diagnostic hysteroscopy procedures. The study took place at the Gynecology and Obstetrics Clinic of Bağcılar Training and Research Hospital, from January 15, 2023, to May 15, 2023.

### 2.2. Participants

A total of 200 female patients, aged 18 years and older, who met the inclusion criteria were enrolled in the study. Eligibility was determined based on the presence of clinical indications for hysteroscopy such as abnormal uterine bleeding, suspected intrauterine lesions, or infertility evaluation. Exclusion criteria included patients with previous cervical surgeries or conditions that could potentially affect the hysteroscopy outcomes.

### 2.3. Data Collection

The hysteroscopies were performed by an experienced gynecologist specializing in minimally invasive gynecological procedures. No speculum, cervical dilation, or anesthesia was used during the procedures to maintain consistency and to highlight the minimally invasive nature of the technique. Both diagnostic (assessment only) and operative (treatment intervention) hysteroscopies were included, with operative procedures involving tasks such as polyp removal or adhesion resolution.

Comprehensive data were gathered on each participant, including demographic information (age, height, weight), reproductive history (gravidity, parity), and details of any past uterine operations. Specific procedural data collected included the indications for hysteroscopy, outcomes of the procedure, pain scores assessed using the Visual Analog Scale (VAS) during cervical canal passage and at the end of the procedure, and total duration of the procedure.

### 2.4. Data Collection

Statistical analyses were performed using SPSS software 25.0. Descriptive statistics summarized the demographics and procedural details. Pain scores and procedural durations were compared between the diagnostic and operative groups using independent t-tests for continuous variables and chi-square tests for categorical variables. Multiple regression analysis was employed to explore the predictors of procedural pain and duration.

This study was approved by the institutional review board (IRB) of Bağcilar Training and Research Hospital. Written informed consent was obtained from all participants before their inclusion in the study. The study was conducted in accordance with the Declaration of Helsinki, and the ethical approval code is (protocol code 2023/03-08 and date of approval February 6, 2023).

All data supporting the findings of this study are available upon reasonable request from the corresponding author. Data related to large datasets have been deposited in a recognized public database, with accession numbers to be provided during the peer review process and prior to publication.

### 2.5. Statistical Analysis

Statistical analyses were conducted using SPSS software, with significance set at p < 0.05. The analyses included chi-square tests for categorical data and t-tests for continuous variables. Results highlighted no significant differences in pain scores, affirming the non-inferiority of operative hysteroscopy in terms of patient discomfort compared to diagnostic procedures.

## 3. Results

This section provides a detailed analysis of the outcomes from the study assessing the efficacy of operative hysteroscopy procedures in comparison to diagnostic hysteroscopy, with specific attention to pain scores and procedural outcomes. The results are presented with subheadings for clarity and include tabulated data for precise interpretation.

### 3.1. Patient Demographics and Baseline Characteristics

Data on patient demographics were collected to ensure comparability between the groups undergoing operative and diagnostic hysteroscopy. Table 1 displays the demographic details.

**Table 1.** Patient demographics and clinical history.

| Variable | Operative HS<br>(n=100) | Diagnostic HS<br>(n=100) | p-value |
| --- | --- | --- | --- |
| Age (years), mean ± SD | 44 ± 10 | 44 ± 10 | 0.986 |
| BMI, mean ± SD | 29 ± 5 | 29 ± 5 | 0.950 |
| Menopause status, n (%) |  |  | 0.728 |
| - Premenopausal | 78 (78%) | 80 (80%) |  |
| - Postmenopausal | 22 (22%) | 20 (20%) |  |
| Previous Uterine Operations, n (%) | 63 (63%) | 59 (59%) | 0.562 |

The mean age and BMI were statistically indistinguishable between the groups, indicating a well-matched cohort. Menopausal status and the prevalence of previous uterine operations were also similar, confirming that the groups were comparable.

### 3.2. Procedural Outcomes

The primary outcomes of the procedures included pain scores assessed by the Visual Analog Scale (VAS) and the duration of the procedures, detailed in Table 2.

**Table 2.**
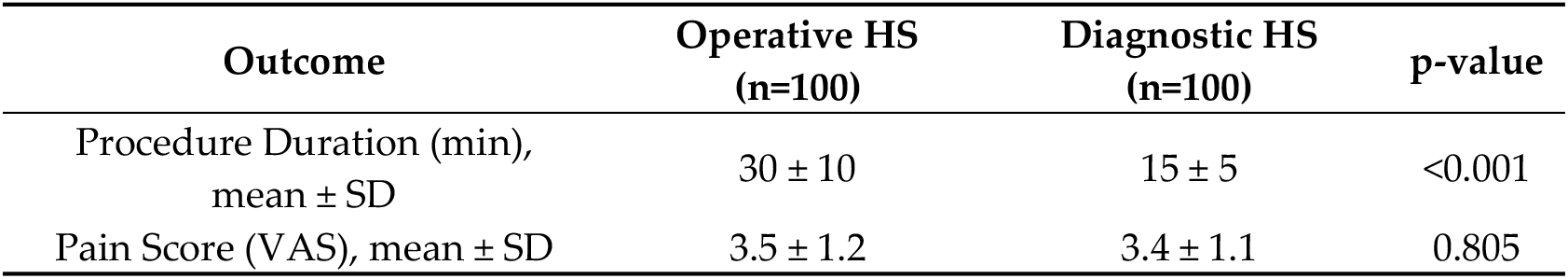
Procedural outcomes and pain assessment.

Operative hysteroscopy procedures were longer, reflecting the complexity and therapeutic nature of these interventions. Pain scores were not significantly different between the groups, suggesting effective pain management regardless of procedure type.

### 3.3. Efficacy of Lesion Removal

The effectiveness of lesion removal during operative hysteroscopy is summarized in Table 3, which categorizes the results by the type of lesion removed.

**Table 3.** Characteristics and Efficacy of Lesion Removal.

| Lesion Type | Completely Removed<br>(n=74) | Not Completely Removed<br>(n=74) | Total |
| --- | --- | --- | --- |
| Polyp | 62 (83.8%) | 12 (16.2%) | 74 |
| Myoma | 2 (2.7%) | 2 (2.7%) | 4 |
| Endometrial<br>Tissue | 9 (12.2%) | 2 (2.7%) | 11 |

Most lesions removed were polyps, with a high success rate of complete removal (83.8%). This emphasizes the effectiveness of operative hysteroscopy in managing common intrauterine pathologies.

## 4. Discussion

The results of this investigation underscore the high efficiency of operative hysteroscopy in the removal of endometrial polyps, achieving an 85.1% complete removal rate. This efficacy is consistent with previous studies, which highlight operative hysteroscopy as a cornerstone procedure for endometrial polyp management due to its minimally invasive nature and high success rates [6,7].

The predominance of polyps among removed lesions (83.8%) observed in this study corresponds with the known epidemiology of endometrial polyps in the gynecological population [8,9]. The successful management of these polyps through operative hysteroscopy not only alleviates symptomatic abnormal uterine bleeding but also may have implications for fertility enhancement, aligning with findings from Al Chami & Saridogan [13] that suggest the removal of polyps could improve reproductive outcomes. Notably, the lack of significant differences in pain scores between the operative and diagnostic groups challenges the notion that more invasive procedures necessarily result in higher pain levels. This finding supports the advancements in surgical techniques and patient care protocols that prioritize minimizing patient discomfort [10,11]. Moreover, this study’s approach to pain management during hysteroscopy—avoiding general anesthesia and using minimal cervical manipulation—could serve as a model for reducing procedural pain across gynecological surgeries.

The minimal impact of menopausal status on pain scores is particularly reassuring, indicating that operative hysteroscopy is a viable option across different patient age groups, including those who are postmenopausal. However, the noted increase in procedural pain among menopausal patients suggests a potential area for further refinement of pain management strategies, possibly incorporating localized analgesic techniques or pre-procedural counseling [12].

Given the association between polyp removal and enhanced fertility outcomes, as highlighted in previous research [13,14], this study further corroborates the importance of incorporating hysteroscopic polypectomy into the fertility-enhancing protocols, especially for patients with unexplained infertility or recurrent implantation failure in assisted reproductive technologies.

The incomplete removal of some polyps, particularly larger or more fibrotic ones, underscores a limitation inherent in the current hysteroscopic techniques. This finding highlights the necessity for ongoing technological and procedural developments, potentially through the adoption of new hysteroscopic instruments or enhanced imaging techniques during surgery [15-17].

From an economic perspective, the ability to perform these procedures in an office setting without anesthesia or extensive preparatory requirements offers significant cost benefits and reduces the patient’s time spent in medical facilities [18-21]. These factors make operative hysteroscopy not only a clinically effective option but also a cost-effective strategy in the management of gynecological pathologies.

Future research should focus on longitudinal studies to assess the recurrence rates of polyps post-hysteroscopy and evaluate the long-term impact on menstrual patterns and fertility. Additionally, comparative studies on different pain management strategies could provide deeper insights into optimizing patient comfort and procedure tolerance.

## 5. Conclusions

This study has validated the effectiveness and safety of operative hysteroscopy for the removal of endometrial polyps, demonstrating a high success rate of 85.1% for complete removal. Key conclusions from the research include:

### High Efficacy

Operative hysteroscopy proves to be a highly effective method for the removal of endometrial polyps, aligning with prior research that emphasizes its role in managing benign gynecological lesions.

### Patient Comfort

The procedure is well-tolerated, with no significant differences in pain scores between patients undergoing operative versus diagnostic hysteroscopy. This underscores the advancements in technique that have minimized patient discomfort.

### Impact on Fertility

The findings suggest that hysteroscopic polypectomy could enhance fertility outcomes, particularly in patients with unexplained subfertility, reinforcing the procedure’s value in fertility treatment protocols.

### Applicability Across Patient Demographics

Menopausal status did not significantly affect pain scores, indicating that operative hysteroscopy is suitable for a broad range of patients, including those who are postmenopausal.

### Need for Continued Innovation

Despite high success rates, the challenge of incomplete removal in some cases highlights the need for continued technological and procedural advancements to improve outcomes.

### Economic and Clinical Advantages

The ability to perform operative hysteroscopy in an office setting offers significant economic benefits and enhances patient convenience, making it a cost-effective approach for the management of gynecological conditions.

In conclusion, operative hysteroscopy is a cornerstone technique in gynecological practice, offering a safe, effective, and patient-friendly approach for the management of endometrial polyps. These results advocate for its continued use and further integration into clinical guidelines, with ongoing research needed to optimize techniques and expand its applicability.

## Data Availability

All relevant data are within the manuscript. Additional anonymized data supporting the findings of this study are available from the corresponding author upon reasonable request.

## Author Contributions

“Conceptualization,T.B. methodology,T.B. software, T.B.; validation,T.B. formal analysis, T.B.; investigation,T.B; resources, T.B.; data curation,T.B. writing—original draft preparation, T.B.; writing—review and editing,T.B.; visualization,T.B. supervision, T.K. project administration, T.B. All authors have read and agreed to the published version of the manuscript.”

## Funding

“This research received no external funding”.

## Institutional Review Board Statement

“The study was conducted according to the Declaration of Helsinki and approved by the Institutional Review Board of Bağcılar Training and Research Hospital (protocol code 2023/03-08 and date of approval February 6, 2023).”

## Conflicts of Interest

“The authors declare no conflicts of interest.”

## Notes

### Competing Interest Statement

The authors have declared no competing interest.

### Funding Statement

This research received no external funding. The study was conducted using institutional resources only. The funders had no role in study design, data collection and analysis, decision to publish, or preparation of the manuscript.

### Author Declarations

This study was approved by the Institutional Review Board of Bağcılar Training and Research Hospital (protocol code: 2023/03-08, approval date: February 6, 2023). Written informed consent was obtained from all participants prior to inclusion in the study. The study was conducted in accordance with the Declaration of Helsinki.

